# BurnCare: Deep Learning–Based Burn Segmentation and Total Body Surface Area Estimation From Smartphone Photographs

**DOI:** 10.64898/2025.12.24.25337638

**Authors:** Abhishek Durgude, Nishant Soni, Kumar Chaitanya Raghuwanshi, Sarthak Awasthi, Kanika Uniyal, Shivam Yadav, Akshat Kakani, Prakher Kesharwani, Vishal Mago, Madhubari Vathulaya, Neeraj Rao, Debarati Chattopadhyay, Akshay Kapoor, Devesh Bhimsaria

**Author notes:** Corresponding Author: Devesh Bhimsaria, Address: Department of Biosciences and Bioengineering, Indian Institute of Technology Roorkee, Roorkee, Uttarakhand 247667, India., Akshay Kapoor, Address: Burn and Plastic Surgery Department, All India Institute of Medical Sciences, Rishikesh, Uttarakhand 249203, India., ORCID ID: https://orcid.org/0000-0003-4223-9039.

## Abstract

**Background:** Burn injuries disproportionately affect resource-limited settings. India alone reports over seven million burn injuries annually against only 67 dedicated burn centres. Consequently, initial assessments, such as total burn surface area (%TBSA), which directly influence fluid resuscitation and referral decisions, are frequently performed by non-specialist staff, with known inter-observer variability. Existing computer-assisted tools either require manual annotation on a generic 3D body model or estimate %TBSA using an anatomical approximation (the Rule of Nines) rather than from the patient’s own segmented body area.

**Methods:** We developed BurnCare, a deployable artificial intelligence-assisted web and Android-based platform that estimates %TBSA from a single smartphone photograph. The pipeline was developed as a two-stage deep learning system that first isolated the patient from the background (ResNet50 encoder, U-Net decoder) and then segmented the burn skin within that region (DenseNet169 encoder, U-Net decoder), with both stages trained on a prospectively collected, multi-device dataset from a tertiary burn centre in India. The platform supports %TBSA estimation both with scale calibration — using a surgical tape of known length — and without it. %TBSA estimated using BurnCare was compared with independent, blinded estimates from three healthcare professionals for 16 patient photographs, using Pearson’s correlation coefficient, mean absolute error (MAE), root mean squared error (RMSE) and the intraclass correlation coefficient (ICC).

**Results:** The background-removal and burn-segmentation models achieved strong performance (intersection over union 0.9811 and 0.8504, respectively). BurnCare’s automatic %TBSA estimate correlated strongly with the mean of three clinicians’ estimates (r = 0.746, p < 0.001, MAE = 6.29, RMSE = 8.63 percentage points; n = 16), improving to r = 0.853 (MAE = 4.32, RMSE = 6.37) after manual mask correction; the ICC among the three clinicians alone was 0.728.

**Conclusions:** A two-stage artificial intelligence-assisted platform, BurnCare, was developed. The platform supports burn detection and %TBSA using deep learning models trained on real-world patient datasets. The foreground prediction is supported by a hybrid ResNet50-U-Net, while burn skin prediction is supported by a DenseNet169-U-Net. BurnCare’s %TBSA estimates agree with clinical judgement about as closely as clinicians agree with each other, supporting its use as a decision-support and documentation tool for burn triage in resource-limited settings. The dataset collected and the BurnCare platform are available online.

## Background

Burn injuries are a prominent cause of accidental death worldwide, where they have been observed in most cases in developing countries. Most developing countries, including India, face a burden due to limited infrastructure for treating such cases. In India alone, over seven million burn injuries occur every year, but the country has only 67 dedicated burn centres to treat them [1,2]. Given these statistics, it is simply not possible to send every patient to the specialised burn centre. So the system should rely on something else: early, accurate triage to determine whether they need to be sent to specialised burn centres or can be treated safely closer to home at peripheral centres. This triage call depends on how much the patient’s body is burned and how serious the skin and tissue damage has occurred. After identifying those patients’ characteristics, a healthcare professional knows how much fluid to provide and when to refer for surgery. Get them wrong, and everything downstream is off. The trouble is that, in day-to-day practice, both are estimated by whoever happens to be on duty. If that person is the regular burn surgeon, their estimate lies closely with the expert consensus. However, in a country with only 67 burn centres serving nearly 1.4 billion people, the people doing the assessment are far more often a general physician or nurse at a peripheral hospital- and their estimates of the total burn surface area (TBSA) are known to vary widely from one assessor to the next, with real consequences for how much fluid a patient gets and whether they are transferred in time [3–7]. Therefore, the role of artificial intelligence is becoming increasingly important.

Existing computer-assisted tools fall short of this goal in a specific way. Commercial 3D-modelling systems require users to manually locate and outline the burn on a generic body model and manually assign its severity — steps that themselves demand burn-management expertise [8]. A more sophisticated approach will be to utilise the resources available to any healthcare professional nowadays: A mobile phone that takes burn patients’ images and do further analysis [9]

Automated burn assessment from images is not new, but every attempt so far has left a piece of the puzzle unsolved [10]. Some earlier work used feature-based classifiers or SVMs [11–14], and others combined CNNs with SVMs conditioned on body part [15, 16] — but these were built and tested as classification tasks, sorting images into categories like ’needs a graft’ or ’does not’, rather than actually measuring burn area pixel by pixel [17–19]. Segmentation-based studies have done better at tracing the boundary between burned and healthy skin [20–26]. However, almost all of them perform segmentation on the entire photo, without first separating the patient from the background clutter. Later researchers [24, 27] have pointed out that this is exactly why background pixels so often get misclassified as burn. The most ambitious effort to date used DeepLabV3+ on a dataset of nearly 5000 images and achieved strong segmentation results, but it was trained on patients of a single ethnicity, so how well it generalises to other skin tones remains an open question. Moreover, like almost every TBSA study before it, it converted the segmented area into %TBSA using the Rule of Nines, a rough anatomical formula rather than a ratio computed from the patient’s own segmented body area [28]. Put simply, no prior pipeline does both things: strip out the background as a dedicated first step and calculate %TBSA directly from the patient’s segmented burn and body areas in the image, with an optional physical-scale reference for absolute wound size, rather than falling back on an anatomical rule of thumb.

We introduce BurnCare, an AI-assisted platform that directly addresses both gaps. We built a two-stage deep learning pipeline — a background-removal stage that isolates the patient before any burn classification is attempted, followed by a dedicated burn-skin segmentation stage, trained on a prospectively collected, multi-device, multi-skin-tone dataset from a tertiary burn centre in India. We further added %TBSA estimation from the processed photo, which supports fluid resuscitation planning. Our objective is to establish this technical pipeline — background separation, burn segmentation, and %TBSA estimation— as the foundation for a triage-support tool whose accuracy we benchmark directly against experienced clinicians’ estimates on the same images — the comparison that ultimately determines whether such a tool belongs in a peripheral-centre workflow.

### BurnCare platform

BurnCare is an AI-assisted platform available as a web-based tool and an Android application, accessed via a secure connection between the user and the server (https://geninfo2.iitr.ac.in/burn_care_2D). BurnCare is powered by deep learning models that were trained on real-world patient data. To use the BurnCare, a user must be a working healthcare professional with a medical practice certificate. The user must first register on the portal, providing basic information, a medical practice certificate, a username, and a password. The BurnCare administration will review all provided information and decide whether to create the user account or reject it. All the users’ information is stored separately and securely on the server.

BurnCare supports bedside photography and AI analysis. Since BurnCare is an AI-based platform, we need data to improve our model. BurnCare respects all users’ and patients’ privacy and obtains prior user consent for the use of their data. The user will be prompted to provide training data; if so, the server will store the user’s uploaded patient data; otherwise, the data will be deleted from the server within 24 hours. More information about the privacy policy and terms and conditions is available. (https://geninfo2.iitr.ac.in/burn_care_2D/privacy.html) Currently, BurnCare offers burn detection and %TBSA estimation from the patient photographs. BurnCare offers single-image and batch-image analysis: for single images, the user can use BurnCare for burn detection and TBSA estimation, while batch analysis is intended only for burn detection. Such a design has been considered for the future addition of features, such as burn-degree modelling, burn-patient 3D modelling, TBSA estimation from 3D burn patient, treatment recommendations, healing-time progression, and whether the user will need to consult at the burn centre. Figure 1 illustrates the overview of the BurnCare app and web server.

**Figure 1:**
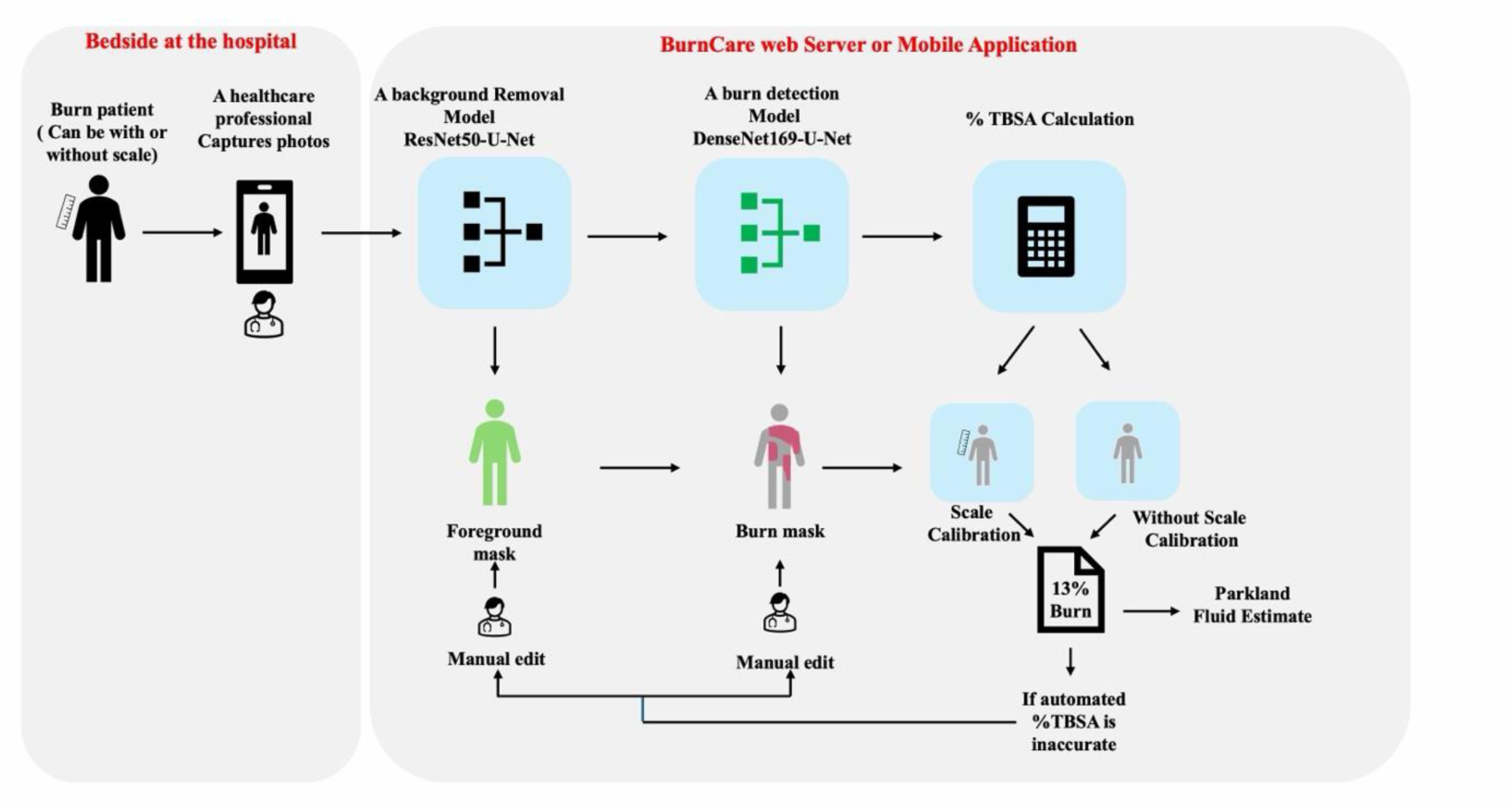
Overview of the BurnCare workflow. A healthcare professional photographs the burn patient at the bedside, with or without a surgical scale reference in frame. The photo is processed by a background-removal model (ResNet50-U-Net) and a burn-detection model (DenseNet169-U-Net) to produce foreground and burn masks, respectively. %TBSA is then calculated, using scale calibration if a reference was included in the photo, or without it otherwise. If the automatic %TBSA estimate is judged inaccurate, the clinician can manually edit the foreground or burn mask, which feeds back into the calculation. The resulting %TBSA is used to calculate a Parkland fluid resuscitation estimate.

## Methods

### Data Collection

Building BurnCare began by collecting real clinical images rather than a curated benchmark dataset. We developed a web-based platform, Care Link (https://geninfo.iitr.ac.in/carelink/login), for this purpose, using PHP. Healthcare professionals in the Burns and Plastic Surgery Department at AIIMS Rishikesh photographed burn patients on their smartphones. They uploaded the images to the Care Link platform, with consent obtained from each patient and, if applicable, their guardian. None of the analysis touched identifiable personal or clinical details — those were left out entirely. By the end of data collection, the total dataset collected was 1003 images from 36 patients at the centre. All the patients were mainly from the state of Uttarakhand, India. Images that were blurry or showed a wound already grafted with tissue were excluded, leaving 803 images for training the BurnCare background-removal stage. All the images were collected after 24-48 hours of the burn injury.

### Stage 1: Background removal

#### Labelling the Data

The first thing BurnCare has to do with any photograph is determine which pixels belong to the patient and which belong to everything else. We treated this as a pixel-wise binary segmentation problem and labelled the training data using prompt-based annotation with the SAM2 model [29], the successor to the original Segment Anything framework [30]. SAM2 is built for exactly this kind of task — it takes a small set of point prompts and returns a precise object mask, making it well-suited to delineating burn patients against cluttered, uncontrolled hospital backgrounds. For each image, we manually placed two to four positive points in the foreground (on the patient) and one or two negative points in the background, and SAM2 returned a binary mask — foreground white, background black. All 803 images that passed quality filtering were labelled this way. Figure 2 shows some representative labelled images.

**Figure 2:**
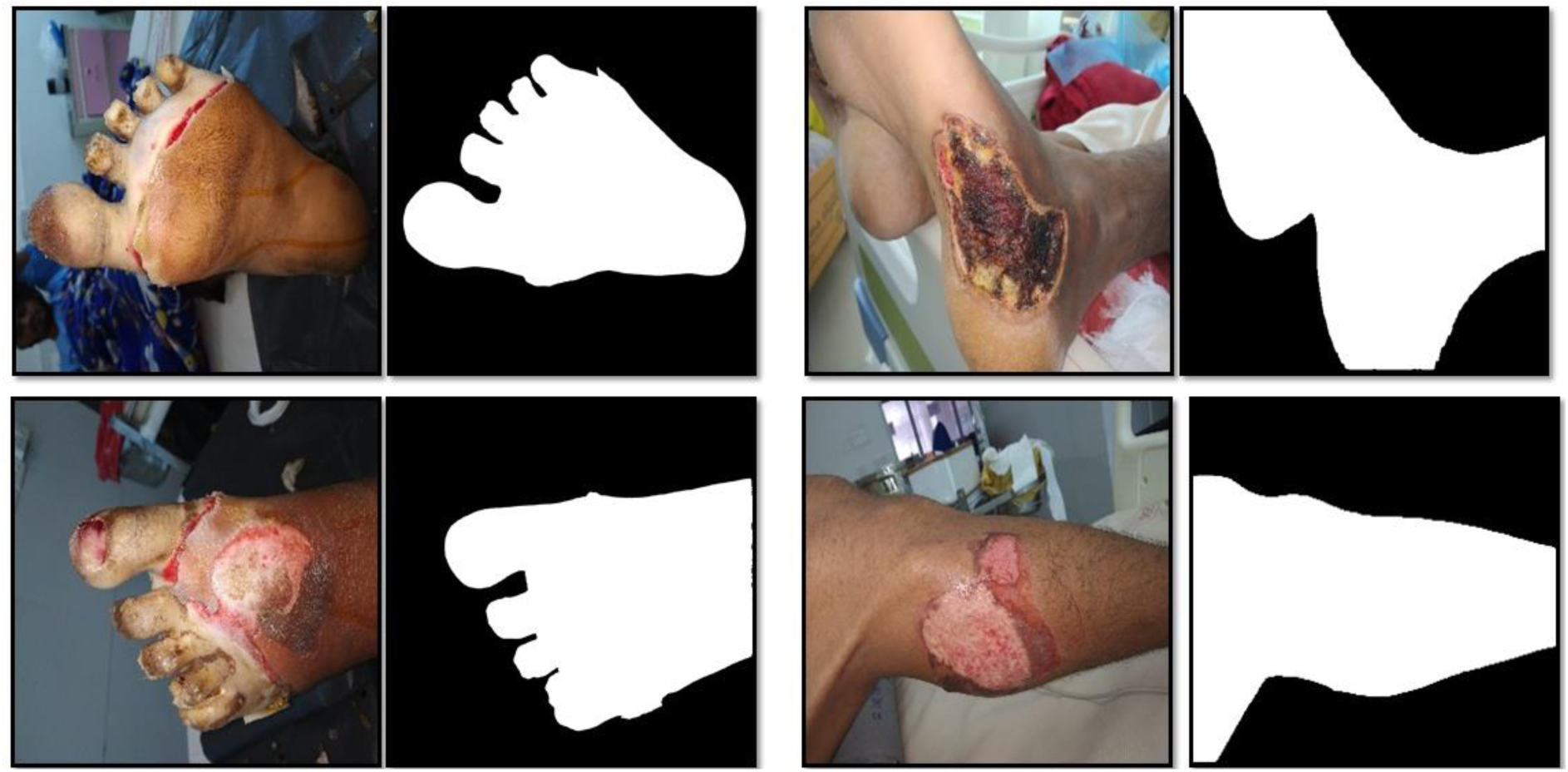
Patients’ photographs, labelled as binary images for background-removal model training. The foreground mask representing the patient’s body is shown as a white mask, and the background is represented as black.

#### Preprocessing and augmentation

To reduce overfitting during model training and help the model generalise better on the unseen dataset, data augmentation was applied. A total of six augmentation approaches (random flip, 90-degree rotation, zoom in, zoom out, transposition and random noise addition) were applied to each image and its corresponding mask identically. Real-world conditions exhibit varied lighting, which we account for in our model when applying the lighting augmentation, including random adjustments to image brightness and contrast. Although applying data augmentation helps mitigate the problem of a limited dataset, it introduces class imbalance in the segmentation problem; therefore, we applied the augmentation to only the training dataset, keeping the validation and test dataset images in their original state. All the images and their corresponding mask were resized to 512 * 512 pixels and each pixel was normalised to the 0-1 range before training.

#### Model Architecture and Training

Convolutional neural networks (CNNs) are a type of deep learning model that help process grid-like data, such as images. The most common use of this network nowadays has been explored in areas such as image segmentation, object detection, scene understanding, and many more. However, when these models are trained from scratch on a small dataset, they tend to overfit, memorising the training data, which in turn results in inaccurate segmentation for new burn patients. Pretrained models help in such cases. These models are trained on the ImageNet dataset, which has already learned about the human body and many other hierarchical visual features, so very little about the burn needs to be learned for such an architecture [31]. Therefore, the model can easily adapt to a smaller number of burn-specific images. At BurnCare, we adopted the hybrid U-Net [32], a CNN model originally developed for medical image segmentation and extensively tested in various image datasets. U-Net has since been paired with transformer encoders [33], reviewed widely as a backbone for biomedical segmentation [34, 35], and extended with attention, residual and nested designs for denoising, lung tumours, cardiac MRI and the pancreas [36–41]. Image segmentation has also been applied to skin lesions [42] and, using U-Net with multi-scale attention, to liver CT [43]; U-Net has further been adapted for unsupervised segmentation [44] and combined with a SAM2 encoder [45]. The model architecture comprised the encoder, which downsamples the image and extracts hierarchical visual features, and the decoder, which restores the spatial resolution lost during encoder downsampling. The U-Net encoder was replaced with a pretrained encoder, such as Visual Geometry Group (VGG) VGG16, VGG19 [46], ResNet50 and ResNet101 [47], all of them trained on the ImageNet dataset. In ResNet variants, residual connections help preserve spatial and contextual details as the network goes deeper. While VGG variants use stacks of convolutional blocks and do not use any residual connections, they are prone to the vanishing gradient problem that commonly occurs in deep networks. The intermediate layers used the rectified linear unit (ReLU) activation function, while the final output layer uses the sigmoid activation function for producing the binary segmentation output. Figure 3 shows the architecture of the hybrid ResNet50–U-Net model.

**Figure 3:**
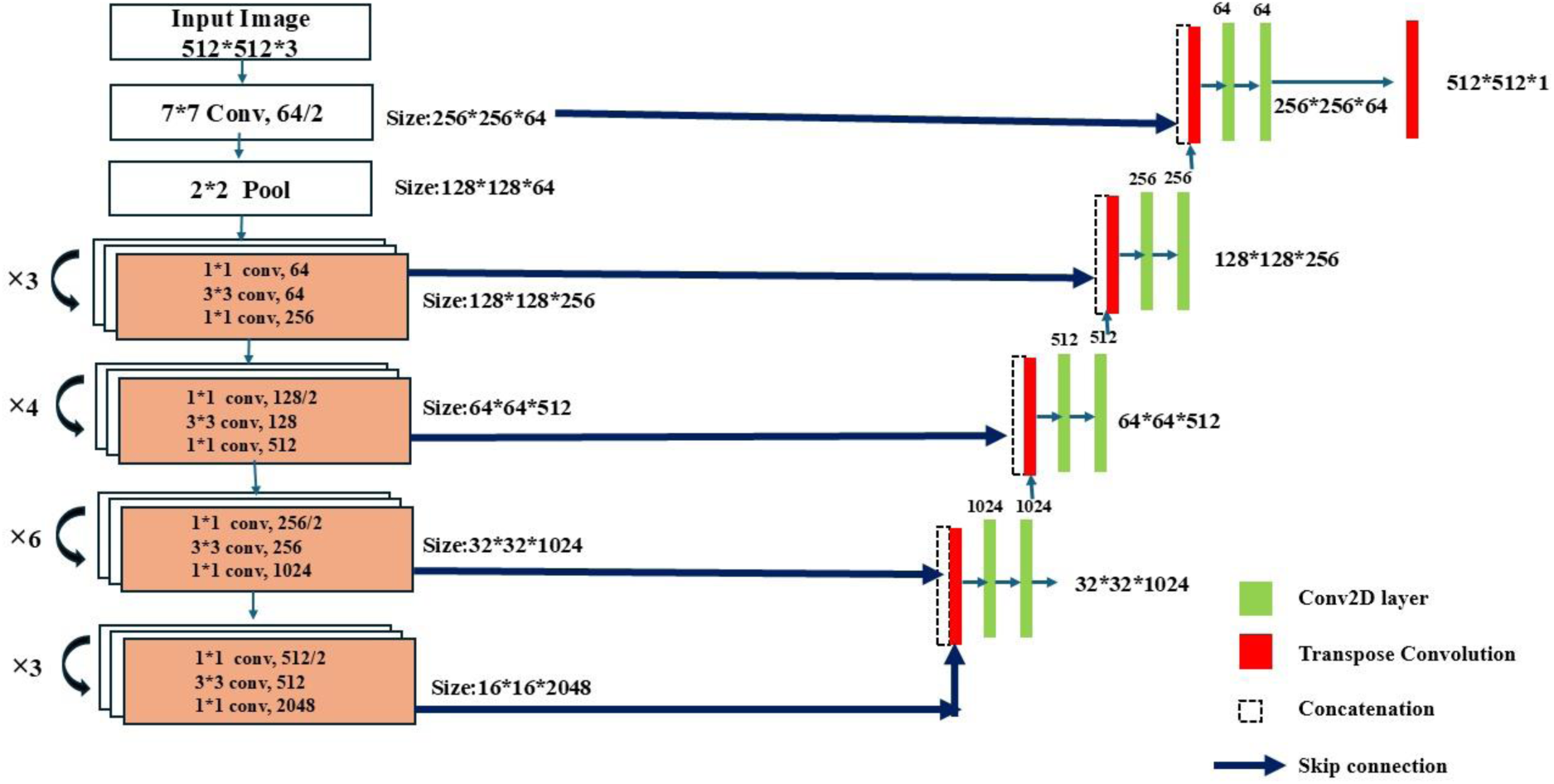
Architecture of hybrid ResNet50-U-Net model used for background removal stage.

Each encoder configuration was trained for 50 epochs with a batch size of 16, the Adam optimizer, and binary cross-entropy loss. A Model Checkpoint callback tracked validation IoU and saved only the best-performing checkpoint.

ResNet50 was selected as the encoder for the background removal task in BurnCare. The model was trained only to isolate the patient’s skin (including both normal and burn skin), hence its output is deliberately restricted to exposed skin rather than the patient’s full silhouette, such as clothing fabric, which can carry textures and colours similar to the burn skin, so classifying burns from the whole photo risks misreading a patterned shirt or a red blanket as a wound. By first isolating only visible skin, this mask ensures the burn-classification stage (Stage 2) never has to make that distinction on clothing at all — it simply never sees it. This skin-only restriction, however, makes the same mask unsuitable as the denominator for %TBSA: a patient wearing clothing over part of their body would have that region silently excluded from the calculation, inflating the reported percentage. BurnCare addresses this with a second, independent mask from Segment Anything Model 2 (SAM2), prompted with a bounding box derived from the ResNet50 skin mask, which segments the patient’s full body, including clothed regions. This whole-body mask, not the skin-only mask, is what BurnCare uses as the %TBSA denominator, while the skin-only mask continues to define where Stage 2 burns are assessed. The two masks answer different questions: one indicates where to look for burns, and the other indicates how large the patient actually is — and neither is a substitute for the other.

### Stage 2: Burn detection

#### Labelling the Data

Once the background is removed, BurnCare’s second stage classifies the remaining skin as normal or burned. We drew 441 background-removed images spanning different body parts — hands, legs, abdomen, and others — and labelled them at the pixel level into three classes using the same SAM2 prompting approach as Stage 1: Class 0, background (blue); Class 1, normal skin (green); Class 2, burned skin (red). For each image, two to four positive points were placed within the burn and one or two negative points on normal skin, which SAM2 then converted into a red burn mask (Figure 4). Background pixels were standardised to blue, and any pixel left unlabelled defaulted to green (normal skin), producing a complete three-class mask (Figure 5).

**Figure 4:**
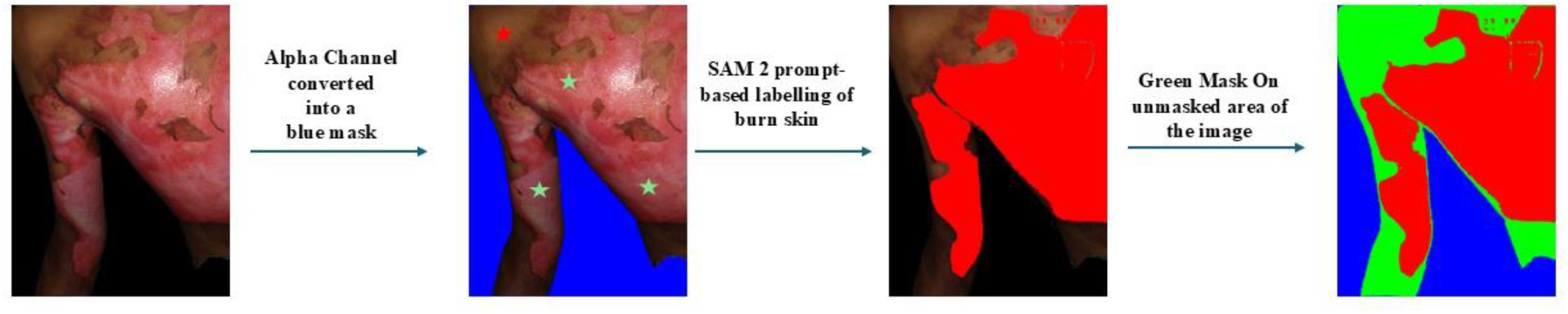
The labelling strategy used to create the training dataset for the burn detection model. SAM2 model is used for burn skin labelling as a red mask. Background and normal skin are shown as a blue and green mask, respectively.

**Figure 5:**
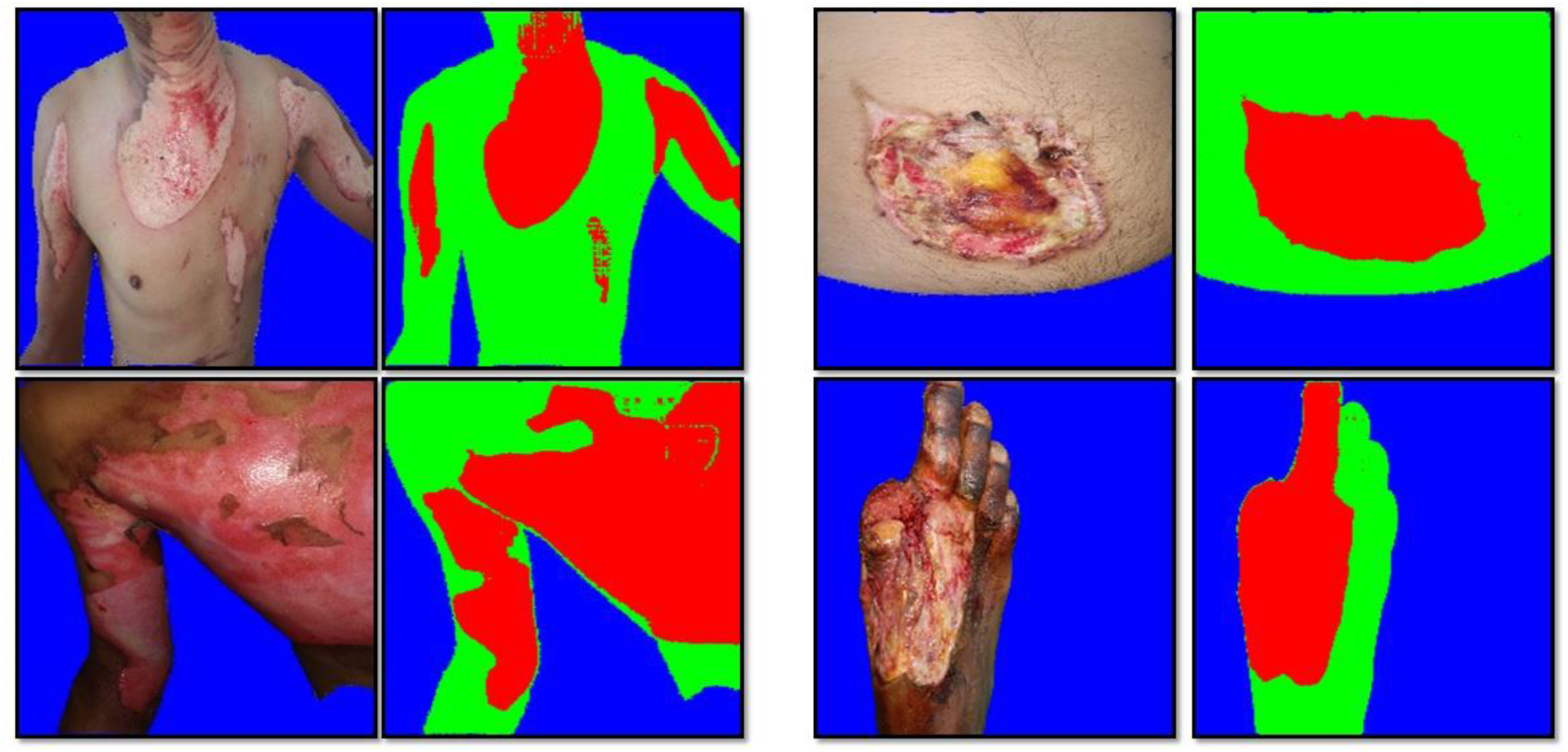
Complete three-class segmentation mask used for burn detection training, with background in blue, normal skin in green and burn skin in red.

#### Class imbalance and loss function

The labelled dataset was imbalanced — 51% background, 27% normal skin, and 21% burn skin pixels — which, left unaddressed, biases a segmentation model toward the majority class. We used weighted categorical cross-entropy to penalise misclassification of normal-skin and burn-skin pixels more heavily than background misclassification.

#### Model Architecture and training

We compared U-Net decoder paired with seven candidate encoders — ResNet50, ResNet101, DenseNet121, DenseNet169 [48], VGG16, VGG19, and EfficientNetB4 [49] — against a baseline U-Net with no pre-trained encoder. Each candidate was trained for 70 epochs, with a batch size of 16, an Adam optimizer, the weighted categorical cross-entropy loss described above, an initial learning rate of 0.00001, and a final 1×1 convolution followed by a softmax activation to produce a three-class output. We tested the model’s performance on held-out images to assess how accurately it differentiated between normal and burned skin. DenseNet169 was selected as the encoder for BurnCare’s deployed burn-detection stage. Rather than running on the full photograph, it processes each connected skin region identified by Stage 1’s mask independently, classifying each into background, normal skin, or burn skin at full resolution, without ever seeing clothed regions of the image.

#### Model Evaluation

Both models for background removal and burn skin detection were evaluated using metrics such as accuracy, Intersection over Union (IoU), precision, recall, and F1-score. All metrics were computed from the model’s predictions of true positives (TP), true negatives (TN), false positives (FP), and false negatives (FN). Models were evaluated at both the training and validation sets, and based on the validation set, the models were tested and used in the backend of the BurnCare. All models were trained using the above metrics in 10-fold cross-validation, and for each architecture, all reported metrics are taken from the best-performing fold. The detailed description of each metric is given below as

Accuracy: Accuracy represents the ratio of correctly classified pixels, including true positives and negatives, to the overall pixel count.

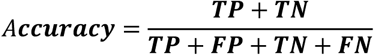

Intersection over union: It measures how much the predicted segmentation overlaps with the ground truth by calculating the ratio of their intersection to their union. A higher IoU means better segmentation accuracy.

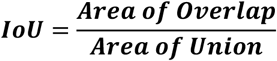

Precision (Positive Predictive Value): A metric that calculates the accuracy of the positive predictions given by the model.

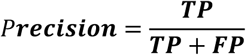

Recall (Sensitivity or True Positive Rate): This metric measures the proportion of relevant instances (true positives) in the data.

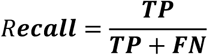

F1-score: The F1-score combines precision and recall into a single metric, providing a balanced measure of both.

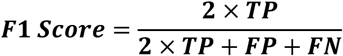

#### Implementation Detail

All the models were trained on a Dell Precision tower 5860 with an Intel® Xeon® w7-2495X (45 MB cache, 24 cores, 2.5 GHz to 4.8 GHz) processor, NVIDIA® RTX™ A6000, 48 GB GDDR6 GPU, and 256 GB RAM

#### Total burn surface area (TBSA) estimation

%TBSA can be calculated with or without a scale reference. When the absolute wound area (cm²) is also required, the clinician photographs the burn with a measuring reference — a ruler or a strip of surgical tape of known length, typically 15 cm — placed visibly in the same frame as the wound. This is the same photograph that will be further processed through background removal (Stage 1) and burn detection (Stage 2).

TBSA is calculated as the fraction of the patient’s body surface area that is burned. Since a single 2D photograph can only capture one side of the patient, BurnCare approximates the unseen side of the body by doubling the whole-body foreground pixel count from the SAM2 mask before using it as the denominator:

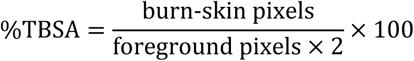

The foreground pixels are the whole-body mask described in Stage 1, derived from the SAM2 model; doubling this value accounts for the body surface area not visible in a single photograph.

The %TBSA is calculated based on the patient’s body seen in the photograph. BurnCare offers TBSA calculation in two ways: with the scale reference shown in the image and without it. For an image with a surgical scale as reference, a percentage alone does not indicate a wound’s actual size. For that, BurnCare needs to convert pixels to centimetres, using the ruler or tape already in the photo: the clinician taps the two ends of the ruler on the screen in the app’s or web server TBSA Calculator and enters its real-world length (15 cm by default, editable if a different reference was used). From the on-screen distance between those two taps and the entered real length, the app derives a pixel-to-centimetre scale and converts the burn area from pixels² to cm². If no ruler or tape is captured in the photo, the clinician can select ’Calculate Without Scale’ — %TBSA is still reported, but no cm² figure is provided.

The TBSA estimation is sensitive to the angle and distance from which the photo is taken, and is influenced by the surgical dressing and clothing. The model never knows what is beneath the clothing, so we considered involving a human in the loop to take action in such cases. We allow users to edit the detected foreground and burn masks after the AI processing. If the user needs to edit the mask, they can add or delete it by selecting the polygon. Once the polygon is saved, the new updated mask is considered for further processing.

To evaluate BurnCare’s %TBSA output against clinical judgement rather than only against another algorithm, we compared it to independent %TBSA estimates from three healthcare professionals at our centre (Clinician 1, Clinician 2, and Clinician 3). Sixteen unique photographs were sent for review. Each clinician viewed the photographs independently, was blinded to BurnCare’s output, and was instructed to estimate %TBSA based only on the burned skin visible in each photograph. We compared BurnCare’s automatic estimate, and separately its estimate after manual mask correction, against the mean of the three clinicians’ estimates using Pearson’s correlation coefficient (r), mean absolute error (MAE), mean squared error (MSE) and root mean squared error (RMSE); MAE and RMSE are expressed in percentage points and MSE in squared percentage points. We separately computed the pairwise Pearson correlation and the same error metrics between each pair of clinicians, and the intraclass correlation coefficient (ICC, two-way random-effects, absolute agreement, single rater) among the three clinicians alone, as a benchmark of ordinary inter-observer variability in this task.

The TBSA is required to estimate fluid resuscitation. BurnCare automatically computes the fluid required using the Parkland formula (4 mL × body weight in kg × %TBSA) given over 24 hours, half in the first 8 hours, using the patient’s recorded weight, and shows it alongside the TBSA result. This turns a segmentation output into an immediately actionable resuscitation volume. This was a proof-of-concept study to assess the feasibility and estimated accuracy of determining TBSA from 2D images. By allowing the human in the loop for clinical assessment, the clinical interpretation from the BurnCare is much more reliable for accurate %TBSA (Figure 6). Readers may contact the corresponding author for Figure 6 since the patient’s full body images, which we can’t disclose publicly.

**Figure 6.** The manual mask-correction workflow in BurnCare’s TBSA Calculator, shown on a real case. On the left, the automatic burn mask indicates 29.38% TBSA, with the entire back and part of the arm marked red. In the middle, the clinician is fixing it — drawing a polygon with the erase tool to pull a patch of overdetected burn back out of the mask, since that first pass included a small quantity of skin it should not have included. On the right, after saving the edit, the mask is tighter, and the estimate has dropped to 28.57% — a small but real correction that came straight from a human catching what the model got slightly wrong.

## Results

### 1) Background Removal

Four pretrained encoders were compared-ResNet50, ResNet101, VGG16 and VGG19. Each encoder was paired with a U-Net decoder. The four-hybrid architecture was trained on burn patients to detect the foreground. The models were evaluated using cross-validation on the 803 background removal dataset. Table 1 summarises their performance.

**Table 1.** Background removal performance by encoder architecture.

| Sr. No. | Models | Accuracy | IoU | Precision | Recall | F1-score |
| --- | --- | --- | --- | --- | --- | --- |
| 1. | ResNet50 | 0.9914 | 0.9811 | 0.9844 | 0.9966 | 0.9905 |
| 2. | ResNet101 | 0.9733 | 0.9428 | 0.9531 | 0.9885 | 0.9705 |
| 3. | VGG16 | 0.9507 | 0.9011 | 0.9252 | 0.9719 | 0.9480 |
| 4. | VGG19 | 0.9380 | 0.8753 | 0.9017 | 0.9676 | 0.9335 |

In a typical burn-patient photograph, background pixels outnumber the patient’s foreground pixels; therefore, relying solely on accuracy is misleading, since more background pixels will contribute to the overall score. IoU is stricter in this setting: it only credits a model for pixels it predicted correctly on both sides of the foreground and background boundary. On the IoU metrics, ResNet50’s advantage is the largest and clearest, with a score of 0.9811 against ResNet101’s 0.9428.

Precision and recall are real predictors of the model’s performance, as their values will determine the model’s downstream use. A false-positive prediction, which segments some background pixels as skin, can pass non-patient regions to the burn-detection stage and enlarge the bounding box used to prompt SAM2 for the %TBSA denominator. In contrast, a false negative, where part of the patient’s skin is wrongly classified as background, hides those pixels from burn detection, so burns in that region are never counted, and can shrink the SAM2 prompt so that part of the body is excluded from the denominator. ResNet50 has both high precision (0.9844) and recall (0.9966) values out of the four candidates, which shows that the model is not trading the one error type for the other; it is simply making fewer errors of both kinds, which is exactly the property this mask needs before it ever reaches the TBSA calculation. F1-score, as the harmonic mean of the two, summarised that balance in a single number (0.9905) and again put ResNet50 clearly ahead of all candidates.

The ResNet101 trailed ResNet50 across every metric, with an IoU of 0.9428 compared to ResNet50’s 0.9811. This observation shows that, despite being a deeper network, the extra depth in ResNet101 did not yield better segmentation here. In terms of precision, the model scores at 0.9531 with a recall of 0.9885, showing some accumulation of false-positive pixels.

VGG16 outperformed VGG19 on every metric, with an IoU of 0.9011 against the IoU of 0.8753 of VGG19, and an F1-score of 0.9480, higher than the F1-score of 0.9335 of VGG19. This observation shows that the extra layers in VGG19 did not yield better segmentation results, since the model lacks residual connections, unlike those in ResNet variants. This architectural distinction between the ResNet and VGG therefore shows how the residual connections in the ResNet help the model generalise better than VGG variants.

Qualitatively, ResNet50’s predicted masks (Figure 7) captured the visible patient cleanly across a range of poses, lighting conditions, and skin tones, correctly including both normal and burned skin in the foreground, thereby providing Stage 2 with a usable region for classification.

**Figure 7:**
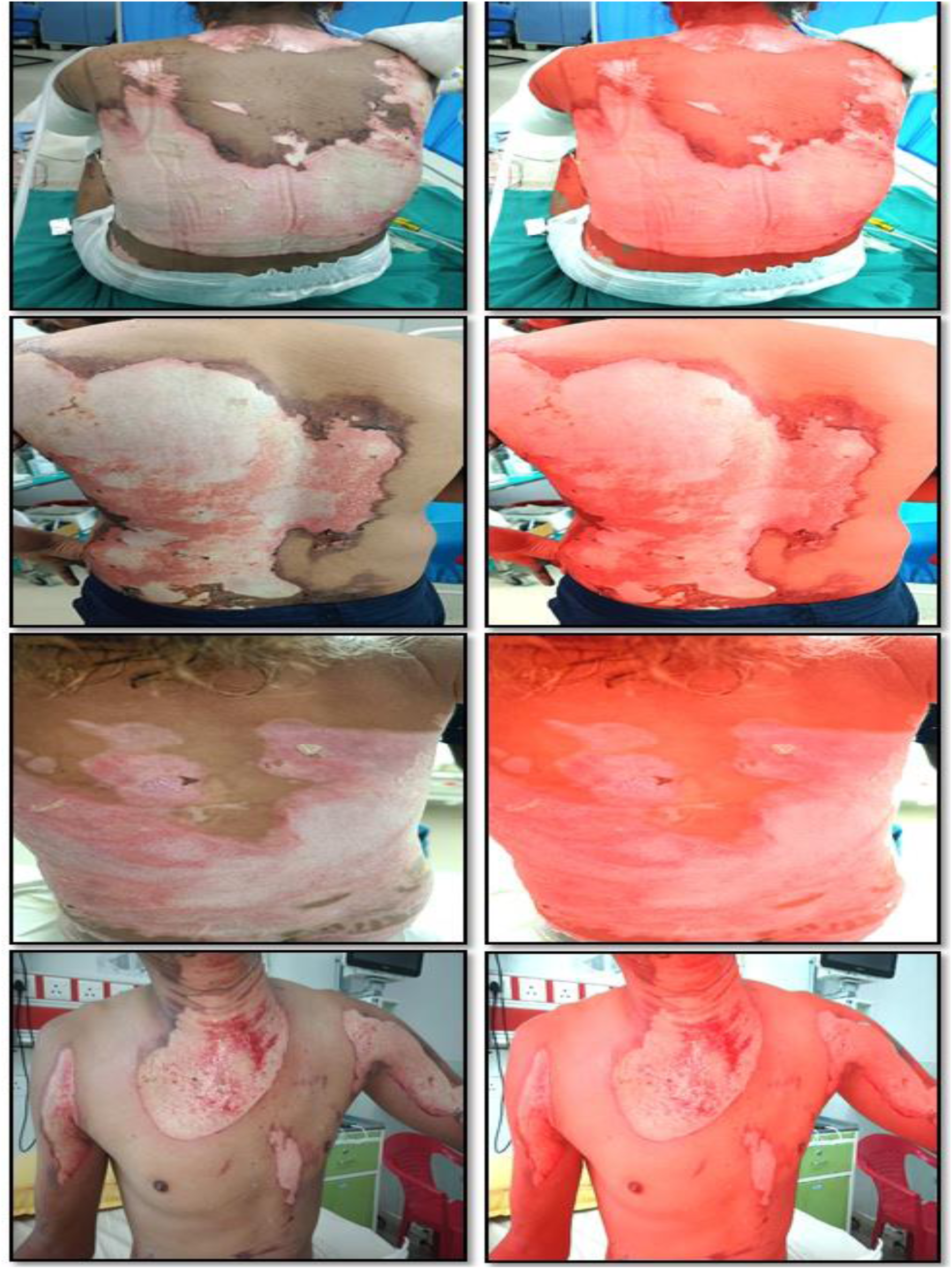
Qualitative result of the ResNet50-U-Net background removal across different body poses and skin tones. Model predicted mask is shown as red mask.

### 2) Burn Detection

Burn detection was performed using the labelled 441-burn image dataset. To carry out the task, we trained seven pre-trained encoders — ResNet50, ResNet101, DenseNet121, DenseNet169, VGG16, VGG19, and EfficientNetB4 — plus a baseline U-Net with no pretrained encoder. Each was evaluated on a three-class segmentation task to detect background, normal skin, and burn skin.

Overall accuracy alone can be misleading in multiclass segmentation tasks and cannot indicate how a model performs on the class that actually matters in clinical decision-making. In such class-imbalance scenarios, per-class precision, recall, and F1-score help determine whether the model is accumulating false positives and false negatives. We observed and reported the class imbalance during model training and took necessary measures to compute metrics accurately.

Background class segmentation is easy for every encoder, as evidenced by high IoU values around 0.98 across all the models. This is expected, since stage 1 has already removed most background pixels before this model ever sees the image, leaving few background pixels for any encoder to get wrong. The classes that are actually separate and relevant to the TBSA calculation are normal skin and burn skin, since burn skin pixels contribute to the TBSA numerator, and errors in these pixels have direct clinical consequences for clinical decisions. ResNet50 and ResNet101 actually had the higher Burn Skin IoU — 0.8583 and 0.8559. Going by that number alone, one of them looks like the better pick. Intersection over Union (IoU) measures the overlap between the ground truth and the model-predicted mask; it does not indicate whether the model can distinguish between normal and burned skin boundaries in an image, nor whether the boundaries are blurred. After we conducted testing on the held-out test data, we observed that ResNet50 and ResNet101 frequently missed the true boundaries between normal and burned skin, even though their IoU scores were slightly higher than those of the other models. DenseNet169 distinguishes boundaries more accurately. The DenseNet architecture consists of dense blocks, in which each layer in the network is connected to all previous layers’ feature maps, not just the feature maps from one previous layer, a design not available in ResNet variants. This dense interconnection helps the model capture the fuzzy, gradual edge of a burn rather than only the bulk of the wound. Since the detected burn pixels are directly used for the %TBSA estimation, getting the edge right matters more than squeezing out a slightly better IoU. DenseNet121 performs slightly worse than DenseNet169 (IoU 0.8490 vs 0.8504), suggesting that the extra depth in DenseNet169 yields a small improvement. Unlike the ResNet variant in stage 1, the VGG variants show poor performance, with VGG16 and VGG19 achieving IoU scores of 0.8427 and 0.8396, respectively. VGG19 particularly shows a large precision-recall gap, with precision and recall of 0.8684 and 0.9619, respectively, indicating the accumulation of false positives in burn skin detection. EfficientNetB4 (IoU 0.8216) and, most clearly, the baseline U-Net with no pre-trained encoder at all (IoU 0.7901) show the worst Burn Skin IoU, as seen in Table 2, which underlines the same point.

**Table 2.** reports accuracy, IoU, precision, recall and F1-score separately for each model for each class.

| Sr. No. | Model Architecture/Metrics | Accuracy |  | Intersection Over Union | Precision | Recall | F1-score |
| --- | --- | --- | --- | --- | --- | --- | --- |
| 1 | ResNet50 (With U-Net Decoder) | 0.9654 | Normal Skin | 0.8889 | 0.9445 | 0.9379 | 0.9412 |
|  |  |  | Burn Skin | 0.8583 | 0.9125 | 0.9352 | 0.9237 |
|  |  |  | Background | 0.9867 | 0.9958 | 0.9908 | 0.9933 |
| 2 | ResNet101(With U-Net Decoder) | 0.9633 | Normal Skin | 0.8581 | 0.9151 | 0.9323 | 0.9237 |
|  |  |  | Burn Skin | 0.8559 | 0.9292 | 0.9156 | 0.9224 |
|  |  |  | Background | 0.9893 | 0.9958 | 0.9934 | 0.9946 |
| 3 | DenseNet121(With U-Net Decoder) | 0.9608 | Normal Skin | 0.8628 | 0.9282 | 0.9245 | 0.9264 |
|  |  |  | Burn Skin | 0.8490 | 0.9056 | 0.9314 | 0.9183 |
|  |  |  | Background | 0.9833 | 0.9954 | 0.9876 | 0.9915 |
| 4 | DenseNet169(With U-Net Decoder) | 0.9662 | Normal Skin | 0.8766 | 0.9240 | 0.9447 | 0.9342 |
|  |  |  | Burn Skin | 0.8504 | 0.9230 | 0.9154 | 0.9192 |
|  |  |  | Background | 0.9873 | 0.9972 | 0.9907 | 0.9936 |
| 5 | VGG16(With U-Net Decoder) | 0.9604 | Normal Skin | 0.8351 | 0.8978 | 0.9229 | 0.9101 |
|  |  |  | Burn Skin | 0.8427 | 0.9286 | 0.9010 | 0.9146 |
|  |  |  | Background | 0.9919 | 0.9956 | 0.9962 | 0.9959 |
| 6 | VGG19(With U-Net Decoder) | 0.9602 | Normal Skin | 0.8548 | 0.8539 | 0.8916 | 0.9217 |
|  |  |  | Burn Skin | 0.8396 | 0.8684 | 0.9619 | 0.9128 |
|  |  |  | Background | 0.9877 | 0.9969 | 0.9908 | 0.9938 |
| 7 | EfficientNetB4(With U-Net Decoder) | 0.9597 | Normal Skin | 0.8524 | 0.9253 | 0.9154 | 0.9203 |
|  |  |  | Burn Skin | 0.8216 | 0.9253 | 0.9064 | 0.9021 |
|  |  |  | Background | 0.9860 | 0.8979 | 0.9939 | 0.9929 |
| 8 | U-Net Only | 0.9601 | Normal Skin | 0.8584 | 0.8841 | 0.9673 | 0.923 |
|  |  |  | Burn Skin | 0.7901 | 0.9528 | 0.8223 | 0.8828 |
|  |  |  | Background | 0.9940 | 0.9972 | 0.9967 | 0.9969 |

Based on both qualitative and quantitative analyses, the deployed DenseNet169 model (Figure 8) accurately identified burned skin across different body parts and correctly traced the boundary between burned and normal skin in most cases.

**Figure 8:**
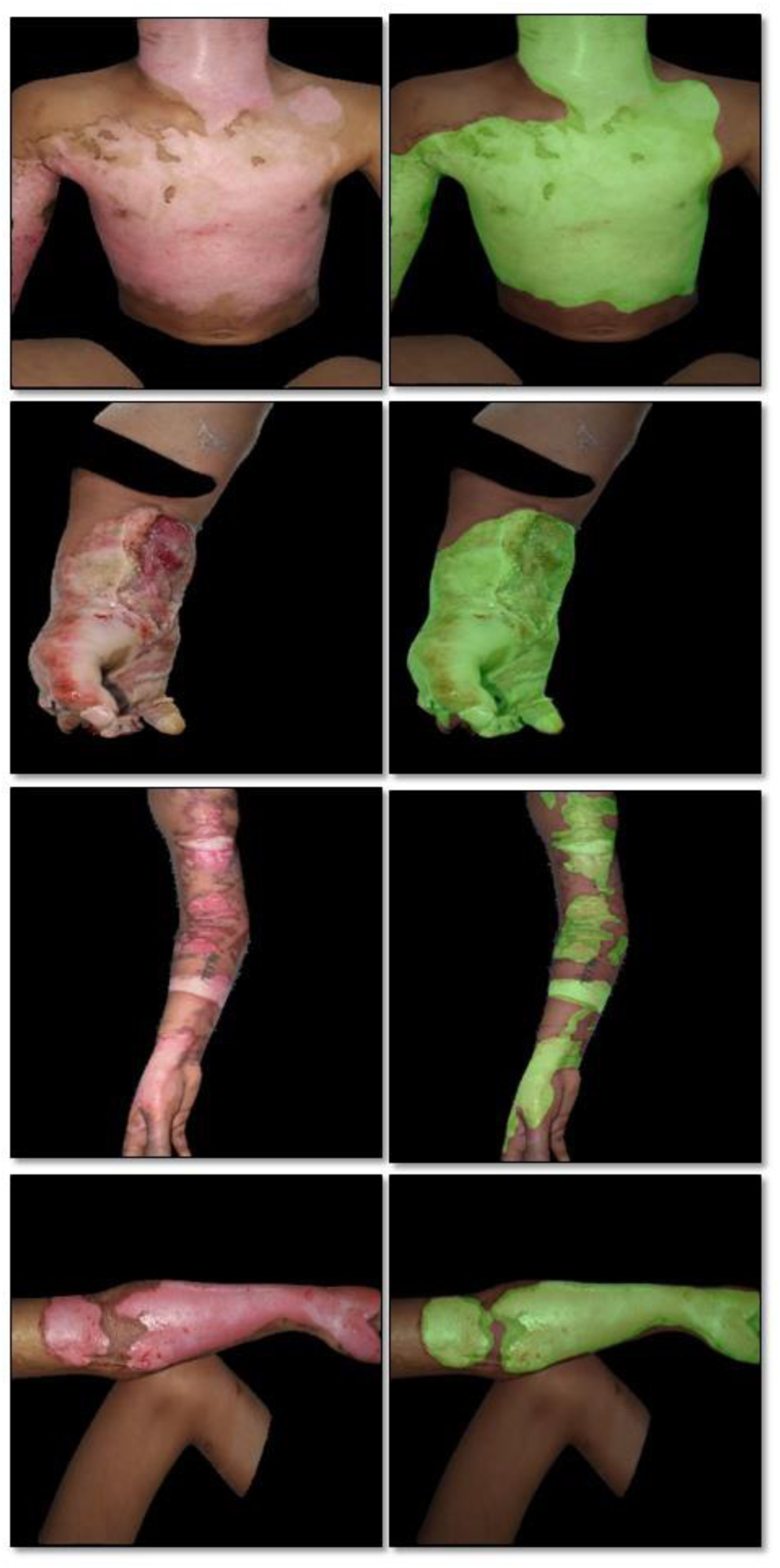
Qualitative DenseNet 169 burn detection result across different body parts. The model prediction of burn skin is shown in the green mask.

#### Total Burn Surface Area Estimation (%TBSA)

**We estimated the %TBSA** from the 2D images and, where a reference scale was present, also determined the wound size in cm². BurnCare supports the %TBSA calculation with and without the scale.

For TBSA estimation with the surgical scale imposed in the image, BurnCare allows the user to draw a line on the surgical scale; the user then has to add the real-world length of that scale. Once the user adds those two, BurnCare automatically calculates the pixel-per-cm conversion factor. This conversion factor converts the foreground mask pixels and burn mask pixels into the cm². Once BurnCare has the foreground and burn masks in cm², it provides the %TBSA value for the given image. Based on the provided output, the user can edit the final mask if needed. When the surgical tape is not present in the burn patient image, BurnCare calculates the %TBSA based on the number of pixels identified for the foreground and burn skin. Figure 9 shows the results of the BurnCare TBSA calculation, with each mathematical step shown.

**Figure 9:**
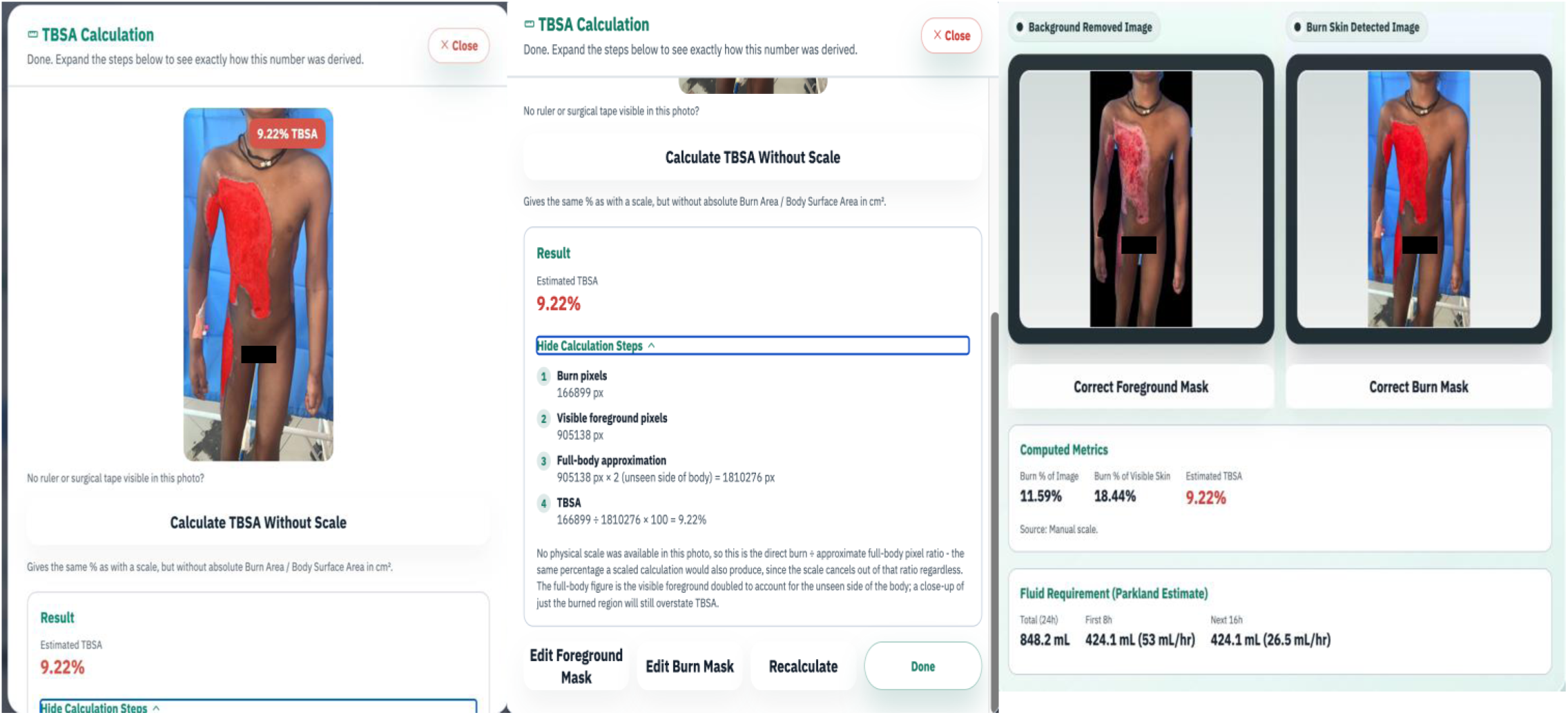
BurnCare’s TBSA Calculator for one example image, showing exactly how the number was worked out, without a scale reference. The app counts burn pixels from the burn-detection mask and whole-body foreground pixels from the SAM2 mask, doubles the foreground count to account for the side of the body the camera can’t see, and divides burn pixels by that doubled figure to get the %TBSA — 9.22% here. Alongside this, the panel shows the background-removed and burn-detected images, the computed metrics (burn as a percentage of the whole image, burn as a percentage of visible skin, and the estimated TBSA), buttons to manually correct either mask if something looks off, and the resulting Parkland fluid-resuscitation estimate.

To assess agreement between BurnCare’s automatic %TBSA estimate and clinical judgement, we compared the model’s output with the three clinicians’ independent estimates across 16 unique photographs. Table 3 lists the per-image comparison.

**Table 3.** Comparative %TBSA analysis: BurnCare versus three clinicians. BurnCare %TBSA (auto) represents the %TBSA given by BurnCare without manual mask intervention, and BurnCare %TBSA (after mask correction) represents %TBSA after manual mask correction. Note:manually corrected mask and derived %TBSA is shown in *

| Patient Photographs | Clinician 1 % | Clinician 2 % | Clinician 3 % | BurnCare %TBSA (Auto) | BurnCare (after mask correction, %) * |
| --- | --- | --- | --- | --- | --- |
| Image 1 | 2 | 5.5 | 4 | 2.61 | 2.61 |
| Image 2 | 40 | 21.5 | 45 | 13.44 | **17.22\*** |
| Image 3 | 40 | 35 | 32 | 34.82 | 34.82 |
| Image 4 | 28 | 32 | 36 | 26.92 | 26.92 |
| Image 5 | 8 | 11.5 | 6 | 7.53 | 7.53 |
| Image 6 | 27 | 24 | 30 | 34.42 | 34.42 |
| Image 7 | 10 | 12 | 15 | 0.43 | **7.09\*** |
| Image 8 | 13.5 | 10 | 15 | 9.22 | 9.22 |
| Image 9 | 41.5 | 29 | 30 | 38.32 | 38.32 |
| Image 10 | 18 | 17 | 20 | 32.46 | **30.13\*** |
| Image 11 | 13.5 | 19 | 30 | 25.31 | 25.31 |
| Image 12 | 37 | 30 | 43 | 26.71 | **36.17\*** |
| Image 13 | 31.5 | 27.5 | 30 | 29.39 | 29.39 |
| Image 14 | 40 | 18 | 36 | 32.14 | **32.86\*** |
| Image 15 | 45 | 40 | 36 | 42.45 | 42.45 |
| Image 16 | 25 | 5 | 27 | 29.95 | **20.01\*** |

**Note:** manually corrected mask and derived %TBSA is shown in \*

BurnCare’s automatic %TBSA estimate correlated strongly with the mean of the three clinicians’ estimates (r = 0.746, p < 0.001, MAE = 6.29, RMSE = 8.63 percentage points; n = 16; Figure 10a), improving to r = 0.853 (p < 0.001, MAE = 4.32, RMSE = 6.37) after manual mask correction (Figure 10b). For comparison, pairwise correlation between the clinicians themselves was r = 0.772 for Clinician 1 vs. Clinician 2 (p < 0.001, MAE = 7.5, RMSE = 9.98; Figure 10c), r = 0.694 for Clinician 2 vs. Clinician 3 (p = 0.003, MAE = 7.88, RMSE = 10.64; Figure 10d) and r = 0.864 for Clinician 1 vs. Clinician 3 (p < 0.001, MAE = 5.44, RMSE = 6.82; Figure 10e) — indicating that BurnCare’s agreement with clinical judgement falls within the same range as ordinary inter-observer variability between clinicians. The intraclass correlation coefficient (ICC) among the three clinicians, computed as a benchmark for ordinary human variability in this task, was 0.728. Table 4 presents the overall analysis comparing BurnCare with all clinicians. After mask correction, BurnCare had the lowest error of any comparison (MSE = 40.58, RMSE = 6.37 percentage points), lower than that of the closest-agreeing clinician pair (Clinician 1 vs. Clinician 3, RMSE = 6.82), and mask correction reduced MSE by 46% (from 74.53). In every comparison, RMSE exceeded MAE by a factor of 1.25–1.47, indicating that a small number of images with large disagreement contributed disproportionately to the overall error.

**Figure 10:**
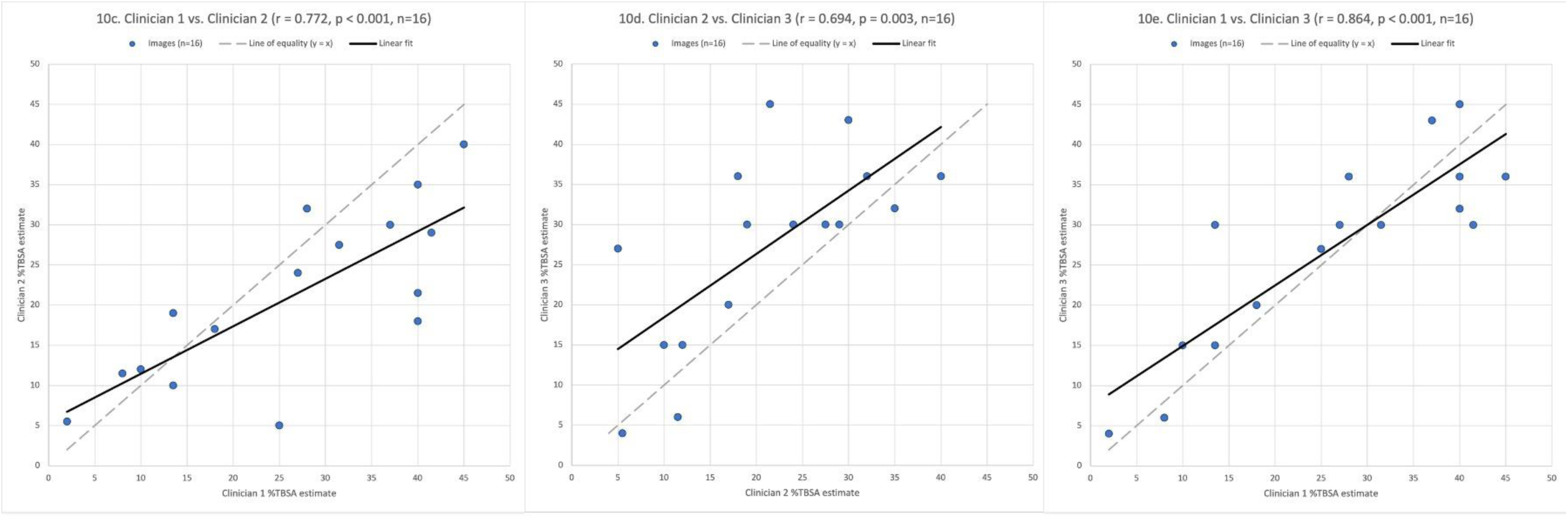
Correlation between BurnCare %TBSA and the mean of the three clinicians’ estimates and Pairwise correlation between the three clinicians’ independent %TBSA estimates (10a) BurnCare %TBSA automatic estimate, without mask correction (r = 0.746, p < 0.001, n = 16).(10b) BurnCare %TBSA estimate, after manual mask correction (r = 0.853, p < 0.001, n = 16). (10c) Clinician 1 vs. Clinician 2 (r = 0.772, p < 0.001). (10d) Clinician 2 vs. Clinician 3 (r = 0.694, p = 0.003). (10e) Clinician 1 vs. Clinician 3 (r = 0.864, p < 0.001). n = 16 for all. Dashed line: perfect agreement (y = x); solid line: linear fit.

**Table 4.** Pearson correlation, mean absolute error, mean squared error and root mean squared error for %TBSA agreement — BurnCare versus clinicians and clinicians versus each other.

| Comparison | Pearson correlation coefficient (r) | Mean absolute error (MAE) | Mean squared error (MSE) | Root mean squared error (RMSE) |
| --- | --- | --- | --- | --- |
| BurnCare %TBSA (Auto) vs. Clinician Mean | 0.746 | 6.29 | 74.53 | 8.63 |
| BurnCare %TBSA (mask Corrected) vs. Clinician Mean | 0.853 | 4.32 | 40.58 | 6.37 |
| Clinician 1 vs Clinician 2 | 0.772 | 7.5 | 99.66 | 9.98 |
| Clinician 2 vs Clinician 3 | 0.694 | 7.88 | 113.13 | 10.64 |
| Clinician 1 vs Clinician 3 | 0.864 | 5.44 | 46.56 | 6.82 |

## Discussion

BurnCare was built to address the specific gaps in the current tools and methods. Current tools rely on manually drawing the burn area on the fixed 3D body model, which requires a person with a strong understanding of anatomy. Most current methods for processing burn patients’ images do not consider background removal. This invites the misclassification of the red blanket, which resembles a similar colour feature to the burn. Since background is an important factor in determining TBSA, which directly depends upon the correct segmentation of the patient’s body, which is only available in the foreground. Incorrect classification, therefore, ultimately propagates into the TBSA calculation, leading to underestimation and overestimation of burn surface area. TBSA has clinical significance during burn treatment planning and fluid resuscitation. Therefore, an error in TBSA estimation can put the patient’s entire life at risk. At BurnCare, we split the problem into sequential steps to process the data. First, the background removal stage removes unnecessary noise, followed by burn segmentation. Both outputs are equally important, as the resulting area will be calculated directly from them. Most of the prior work in this space, including the studies compared above, describes a research pipeline evaluated on a held-out test set and stops there. BurnCare is a deployed system, a web portal and an Android application that healthcare professionals at a tertiary centre can use on real-world patients’ images, extending the mobile-deployment approach explored in prior work [50] with the TBSA output wired directly into a Parkland formula fluid resuscitation calculation, not just as a number the clinician has to act on manually.

The background removal pipeline has a clean takeaway. The ResNet50 performs well on the patient data, achieving the best overlap with the true patient outline with an IoU of 0.9811, precision of 0.9844 and recall of 0.9966. The overall metrics show that the model makes a few mistakes in predicting the foreground. This combination of metrics matters more, since the model-predicted mask defines the region searched for burns in Stage 2 and supplies the bounding-box prompt for the SAM2 whole-body mask used as the %TBSA denominator. A model that misses patient skin can leave burns uncounted and shrink the SAM2 prompt, while a model that incorrectly includes background pixels can introduce false burn detections and enlarge the SAM2 prompt beyond the patient. ResNet50 was more accurate in both cases. VGG variants comparison adds a useful point. VGG19, although a deeper network than VGG16, actually performed worse, with an IoU of 0.8753 compared to VGG16’s 0.9011. More layers in the VGG19 could have helped the model achieve better segmentation, but the model may have been limited by the vanishing gradient problem. That architectural limitation is what the ResNet variant addressed through its residual connection.

For burn segmentation, the IoU numbers alone would have pointed to ResNet50 or ResNet101 over DenseNet169. We did not go with the higher IoU. On held-out images, ResNet50 and ResNet101 both missed the true boundary between normal and burned skin often enough that we did not use them for deployment, even though the overlap metric looked better on paper — IoU rewards getting the bulk of the wound right, not the edge, and the edge is exactly what turns into a clinically meaningful cm² figure. DenseNet169’s dense connectivity, where every layer sees every earlier layer’s features rather than just the one before it, produced visibly cleaner boundaries, and that is why it is the model actually deployed in BurnCare. This is the same reasoning Kuan et al. [24] and Sevik et al. [27] ran into from the other direction — both reported burn/background confusion and patchy, inconsistent masks when segmentation was not preceded by an explicit background-removal step, which is the problem BurnCare’s two-stage design was built specifically to avoid.

Chang et al. [28] remains the closest prior comparison in terms of scale; they collected nearly 5000 images and achieved strong DeepLabV3+ performance, but two things separate BurnCare from it. First, their pipeline didn’t remove the background before segmenting; the same research group has also explored complementary approaches to burn-wound assessment [51, 52]. Also, their models were trained on a single-ethnicity cohort with acknowledged uncertainty about how well their model will generalise to people of different ethnicities; on the other hand, our dataset, collected across a range of skin tones and devices at an Indian tertiary centre, was built with that generalisation question in mind from the start. Second, and more fundamentally, they converted segmented area to %TBSA using the Rule of Nines — the same anatomical approximation the triage literature has been trying to move past [3–7] — while BurnCare computes %TBSA directly from the patient’s segmented burn and whole-body areas in the photo, so the percentage it reports is derived from the patient’s own image rather than an anatomical estimate applied after the fact; a physical scale reference, when present, additionally gives the wound area in cm².

The clinician-comparison results support BurnCare’s automatic %TBSA estimate as a genuine decision-support figure rather than a research-only metric: its correlation with clinical judgement (r = 0.746, improving to r = 0.853 after mask correction) falls within the same range as the clinicians’ own pairwise correlation with one another (r = 0.694–0.864). Its error was likewise within the clinicians’ own range (RMSE = 8.63 percentage points automatically and 6.37 after mask correction, versus 6.82–10.64 between clinician pairs), and the ICC of 0.728 among the clinicians alone indicates the level of inter-observer variability ordinarily expected in this task.

A single-centre sample of this size reflects a real constraint of burn care research in a resource-limited setting, not a shortcut in study design. A larger, ideally multi-centre, prospective comparison remains the clear next step. The planned 3D reconstruction pipeline (see Future Directions), which would remove the single-view full-body approximation that this 2D study relies on, will require the same kind of clinical validation once developed.

### Limitation

Currently, BurnCare models are trained on a single-institute collaboration dataset that includes various body parts and skin tones. Since AI models are very data-hungry, the variation in the dataset and the sheer number of them matter as well. Data augmentation is a strategy to address this issue. Still, it introduces class imbalance in already imbalanced classes, which is the case right now, where the burn area is very small compared to the normal skin and background pixels. In recent days, AI-generated images have helped address the problem of limited datasets, but training models on such datasets and their performance on real patient datasets remain open questions. Secondly, as we described earlier, estimating the %TBSA from 2D images involves assumptions, such as what is present on the other side of the body and whether it’s normal skin or burn skin. Also, patients living in 3D space and their perspective transformations of 2D images do not capture what is present on the curved body surface in burn areas, leading to an underestimation of the burn percentage because fewer pixels cover these areas. Apart from this, a burn patient wearing a dressing and clothes poses one issue: our model never knows what lies beneath the clothing and dressing, whether it’s burn skin or normal skin. Based on this, the %TBSA can be underestimated or overestimated. Keep in mind that we do allow users to provide manual input by editing the mask. We acknowledge two determining factors, namely %TBSA and burn depth, which help greatly in planning a treatment regimen and in determining how long the patient needs to be hospitalised and to recover. Currently, the study does not support burn depth detection; an upcoming investigation will explore the feasibility of burn degree estimation, building on prior burn depth classification work [53, 54].

### Future Directions

BurnCare is designed to be a comprehensive tool that provides all patient information on a single platform. Currently, BurnCare provides AI assistance for burn detection and %TBSA estimation. The next step is burn depth classification. Initial investigation suggests that determining burn depth from RGB images alone is not feasible, as different burn degrees share many similar colour features; multimodal data integration is therefore needed for burn degree classification. In parallel, 3D burn-patient modelling to estimate %TBSA is a planned direction, using multi-stereo vision technology, a parametric model, and a deep learning-based tool. Further collaboration to collect a larger dataset would support research validation and model training. Healing-progression tracking and prediction is a further possible extension: time-series and image data collected over a patient’s treatment course could be used to model expected healing time. Nosocomial infections acquired during a burn patient’s hospital stay, often linked to antimicrobial resistance, present another avenue — bacterial culture data from burn patients, including the antibiotics used and the organisms’ susceptibilities and resistance patterns, could be used to train a model to support antibiotic selection. Each of these directions would need to be investigated and validated against healthcare professionals’ output before deployment.

## Conclusions

Burn care in a country such as India is constrained by a fundamental mismatch: over seven million burn injuries occur annually, yet there are only 67 dedicated burn centres. Hence, a general physician or nurse far more often performs the initial assessment of a burn patient than a burn specialist. BurnCare was developed to address this constraint directly. Rather than requiring a generic 3D body model to be manually annotated by someone with anatomical expertise, the system requires only what a peripheral-centre worker already has access to — a smartphone photograph — and performs patient segmentation, burn detection, and %TBSA estimation from that photograph alone, with optional physical-scale calibration for absolute wound area. The results presented from this study support the feasibility of this approach. For background removal, ResNet50 achieved an IoU of 0.9811 (F1-score 0.9905), and for burn detection, DenseNet169 achieved a burn-skin IoU of 0.8504. BurnCare’s automatic %TBSA estimate correlated strongly with healthcare professional judgement (r = 0.746, RMSE = 8.63 percentage points, improving to r = 0.853, RMSE = 6.37 after mask correction), a level of agreement comparable to the clinicians’ own pairwise agreement with one another (r = 0.694–0.864, RMSE = 6.82–10.64; ICC = 0.728 among the three clinicians). This finding suggests that BurnCare’s disagreement with clinical assessment is not a separate kind of error introduced by the model — it looks more like the same ordinary disagreement you would expect between any two clinicians assessing the same case. Taken together, these findings suggest that BurnCare is a useful decision-support and documentation tool for burn triage in resource-limited settings. Building on this foundation, BurnCare will further investigate and validate the pipeline for 3D burn-patient modelling to estimate %TBSA and classify burn depth.

## Declarations

### Ethics approval and consent to participate

Ethical approval for this study was obtained from the Institutional Ethics Committee of the All India Institute of Medical Sciences (AIIMS), Rishikesh, for the collection and use of data from burn patients. Informed consent was obtained from all patients for the use of their images in this research. The Institutional Ethics Committee of the Indian Institute of Technology (IIT) Roorkee provided provisional ethical clearance for the development and analysis of the artificial intelligence algorithm.

## Consent for publication

All participants included in the study have given their consent prior to the photograph being captured and used for the conduct and publication of the study.

## Availability of data and materials

The annotated data collected during the research is available online for researchers working in the field at https://geninfo.iitr.ac.in/projects, and the webserver Android mobile application is available at https://geninfo2.iitr.ac.in/burn_care_2D/.

## Authors’ contributions

AD and NS worked together on the study’s design and methodology, carried out the investigation, built the software, and handled the formal analysis and data curation; AD also wrote the first draft of the manuscript. CR and SA helped build the software and worked on the analysis and data curation. SY worked on the software and helped with data curation. KU contributed to the software, the analysis, and data curation, and also helped write the first draft. AKa and PK handled data curation. VM took care of visualization, supervision, and project administration. MV, NR, and DC provided resources and oversaw visualization, supervision, and project administration. AKp helped shape the study’s concept and methodology, carried out validation, provided resources, reviewed and edited the writing, and contributed to visualization, supervision, and project administration. DB conceived and designed the study, contributed to the methodology, validation, and investigation, provided resources, reviewed and edited the writing, oversaw visualization, supervision, and project administration, and secured the funding for the work. All authors read and approved the final manuscript.

## Data Availability

All data produced are available online at: https://geninfo.iitr.ac.in/projects

https://geninfo2.iitr.ac.in/burn_care_2D/

## Acknowledgements

We used the artificial intelligence tool Claude (Anthropic) to write the web server and mobile application code.

## References

1. NABI Website. Burn Units and Burn Care Providers in India. Available from: https://thenabi.org/

2. Ahuja RB, Goswami P. Cost of providing inpatient burn care in a tertiary, teaching, hospital of North India. Burns. 2013;39(4):558–64. 10.1016/j.burns.2013.01.013

3. Figueroa C, Palafox MV, Gutierrez EAM, et al. Burn vs. Referral Physicians’ TBSA Estimation Errors: A Cross-Sectional Study at the National Burn Center. Burns Open. 2024;8(4):100361. 10.1016/j.burnso.2024.100361

4. Pham TN, Collier Z, Gillenwater J. Changing the Way We Think About Burn Size Estimation. J Burn Care Res. 2019;40(1):1–11. 10.1093/jbcr/iry050

5. Freiburg C, Igneri P, Sartorelli K, et al. Effects of Differences in Percent Total Body Surface Area Estimation on Fluid Resuscitation of Transferred Burn Patients. J Burn Care Res. 2007;28(1):42–48. 10.1097/bcr.0b013e31802c88b2

6. Sritharan T, et al. Temporal trends in burn size estimation and the impact of the NSW Trauma App on estimation accuracy. Burns. 2023;49(6):1403–11. 10.1016/j.burns.2023.02.002

7. Giretzlehner M, Dirnberger J, Owen R, et al. The determination of total burn surface area: How much difference? Burns. 2013;39(6):1107–13. 10.1016/j.burns.2013.01.021

8. Prieto MF, Acha B, Gómez-Cía T, et al. A system for 3D representation of burns and calculation of burnt skin area. Burns. 2011;37(7):1233–40. 10.1016/j.burns.2011.05.018

9. Xu X, Bu Q, Xie J, et al. On-site burn severity assessment using smartphone-captured color burn wound images. Comput Biol Med. 2024;182. 10.1016/j.compbiomed.2024.109171

10. Huang S, Dang J, Sheckter CC, et al. A systematic review of machine learning and automation in burn wound evaluation: A promising but developing frontier. Burns. 2021;47(8):1691–704. 10.1016/j.burns.2021.07.007

11. Acha B, Serrano C, Acha JI, et al. Segmentation and classification of burn images by color and texture information. J Biomed Opt. 2005;10(3):034014. 10.1117/1.1921227

12. Khani ME, Harris ZB, Osman OB, et al. Supervised machine learning for automatic classification of in vivo scald and contact burn injuries using the terahertz Portable Handheld Spectral Reflection (PHASR) Scanner. Sci Rep. 2022;12(1). 10.1038/s41598-022-08940-4

13. Yadav DP, Sharma A, Singh M, et al. Feature extraction based machine learning for human burn diagnosis from burn images. IEEE J Transl Eng Health Med. 2019;7. 10.1109/JTEHM.2019.2923628

14. Zhang B, Zhou J. Multi-feature representation for burn depth classification via burn images. Artif Intell Med. 2021;118. 10.1016/j.artmed.2021.102128

15. Chauhan J, Goswami R, Goyal P. Using Deep Learning to Classify Burnt Body Parts Images for Better Burns Diagnosis. In: Lepore N, Brieva J, Romero E, Racoceanu D, Joskowicz L, editors. Processing and Analysis of Biomedical Information. SaMBa 2018. Lecture Notes in Computer Science, vol 11379. Cham: Springer; 2019. 10.1007/978-3-030-13835-6_4

16. Chauhan J, Goyal P. BPBSAM: Body part-specific burn severity assessment model. Burns. 2020;46(6):1407–23. 10.1016/j.burns.2020.03.007

17. Boissin C, Laflamme L, Jian F, et al. Development and evaluation of deep learning algorithms for assessment of acute burns and the need for surgery. Sci Rep. 2023;13(1). 10.1038/s41598-023-28164-4

18. Jacobson MJ, Masry M el, Arrubla DC, et al. Autonomous Multi-modality Burn Wound Characterization using Artificial Intelligence. Mil Med. 2023;188:674–81. 10.1093/milmed/usad301

19. Suha SA, Sanam TF. A deep convolutional neural network-based approach for detecting burn severity from skin burn images. Mach Learn Appl. 2022;9:100371. 10.1016/j.mlwa.2022.100371

20. Chauhan J, Goyal P. Convolutional neural network for effective burn region segmentation of color images. Burns. 2021;47(4):854–62. 10.1016/j.burns.2020.08.016

21. Chauhan J, Rosin PL, Goyal P. Burnsnet: burn region segmentation network from color images with two-way CNN. Proc Int Conf Image Process ICIP. 2024:3172–78. 10.1109/ICIP51287.2024.10647789

22. Cirillo MD, Mirdell R, Sjöberg F, et al. Tensor Decomposition for Colour Image Segmentation of Burn Wounds. Sci Rep. 2019;9(1). 10.1038/s41598-019-39782-2

23. Jiao C, Su K, Xie W, et al. Burn image segmentation based on Mask Regions with a Convolutional Neural Network deep learning framework: More accurate and more convenient. Burns Trauma. 2019;7. 10.1186/s41038-018-0137-9

24. Kuan PN, Chua S, Safawi EB, et al. A Comparative Study of Segmentation Algorithms in the Classification of Human Skin Burn Depth. Int J Adv Sci Eng Inf Technol. 2020;10(1):145–50. 10.18517/ijaseit.10.1.10227

25. Liu H, Yue K, Cheng S, et al. A Framework for Automatic Burn Image Segmentation and Burn Depth Diagnosis Using Deep Learning. Comput Math Methods Med. 2021;2021. 10.1155/2021/5514224

26. Rangel-Olvera B, Rosas-Romero R. Detection and classification of skin burns on color images using multi-resolution clustering and the classification of reduced feature subsets. Multimed Tools Appl. 2024;83(18):54925–49. 10.1007/s11042-023-17550-9

27. Şevik U, Karakullukçu E, Berber T, et al. Automatic classification of skin burn colour images using texture-based feature extraction. IET Image Process. 2019;13(11):2018–28. 10.1049/iet-ipr.2018.5899

28. Chang CW, Ho CY, Lai F, et al. Application of multiple deep learning models for automatic burn wound assessment. Burns. 2023;49(5):1039–51. 10.1016/j.burns.2022.07.006

29. Ravi N, Gabeur V, Hu YT, et al. SAM 2: Segment Anything in Images and Videos. 2024. http://arxiv.org/abs/2408.00714

30. Kirillov A, Mintun E, Ravi N, et al. Segment Anything. 2023. http://arxiv.org/abs/2304.02643

31. Rodrigues D de A, Ivo RF, Satapathy SC, et al. A new approach for classification skin lesion based on transfer learning, deep learning, and IoT system. Pattern Recognit Lett. 2020;136:8–15. 10.1016/j.patrec.2020.05.019

32. Ronneberger O, Fischer P, Brox T. U-Net: Convolutional Networks for Biomedical Image Segmentation. 2015. http://arxiv.org/abs/1505.04597

33. Chen J, Lu Y, Yu Q, et al. TransUNet: Transformers Make Strong Encoders for Medical Image Segmentation. 2021. http://arxiv.org/abs/2102.04306

34. Hephzibah R, Anandharaj HC, Kowsalya G, et al. Review on Deep Learning Methodologies in Medical Image Restoration and Segmentation. Curr Med Imaging. 2022;19(8). 10.2174/1573405618666220407112825

35. Rizwan I Haque I, Neubert J. Deep learning approaches to biomedical image segmentation. Inform Med Unlocked. 2020;18. 10.1016/j.imu.2020.100297

36. Rajamani KT, Rani P, Siebert H, et al. Attention-augmented U-Net (AA-U-Net) for semantic segmentation. Signal Image Video Process. 2023;17(4):981–89. 10.1007/s11760-022-02302-3

37. Zhang H, Lian Q, Zhao J, et al. RatUNet: Residual U-Net based on attention mechanism for image denoising. PeerJ Comput Sci. 2022;8. 10.7717/peerj-cs.970

38. Zhou T, Dong Y, Lu H, et al. APU-Net: An Attention Mechanism Parallel U-Net for Lung Tumor Segmentation. BioMed Res Int. 2022;2022. 10.1155/2022/5303651

39. Zhou Z, Siddiquee MMR, Tajbakhsh N, et al. UNet++: A Nested U-Net Architecture for Medical Image Segmentation. 2018. http://arxiv.org/abs/1807.10165

40. Li YZ, Wang Y, Huang YH, et al. RSU-Net: U-net based on residual and self-attention mechanism in the segmentation of cardiac magnetic resonance images. Comput Methods Programs Biomed. 2023;231. 10.1016/j.cmpb.2023.107437

41. Oktay O, Schlemper J, Folgoc L le, et al. Attention U-Net: Learning Where to Look for the Pancreas. 2018. http://arxiv.org/abs/1804.03999

42. Sumithra R, Suhil M, Guru DS. Segmentation and classification of skin lesions for disease diagnosis. Procedia Comput Sci. 2015;45(C):76–85. 10.1016/j.procs.2015.03.090

43. Wu J, Zhou S, Zuo S, et al. U-Net combined with multi-scale attention mechanism for liver segmentation in CT images. BMC Med Inform Decis Mak. 2021;21(1). 10.1186/s12911-021-01649-w

44. Xia X, Kulis B. W-Net: A Deep Model for Fully Unsupervised Image Segmentation. 2017. http://arxiv.org/abs/1711.08506

45. Xiong X, Wu Z, Tan S, et al. SAM2-UNet: Segment Anything 2 Makes Strong Encoder for Natural and Medical Image Segmentation. 2024. http://arxiv.org/abs/2408.08870

46. Simonyan K, Zisserman A. Very deep convolutional networks for large-scale image recognition. Proc Int Conf Learn Represent (ICLR). 2015. 10.48550/arXiv.1409.1556

47. He K, Zhang X, Ren S, et al. Deep residual learning for image recognition. Proc IEEE Conf Comput Vis Pattern Recognit (CVPR). 2016. 10.48550/arXiv.1512.03385

48. Huang G, Liu Z, van der Maaten L, et al. Densely connected convolutional networks. Proc IEEE Conf Comput Vis Pattern Recognit (CVPR). 2017. 10.48550/arXiv.1608.06993

49. Tan M, Le QV. EfficientNet: Rethinking model scaling for convolutional neural networks. Proc 36th Int Conf Mach Learn (ICML). 2019. 10.48550/arXiv.1905.11946

50. Yıldız M, Sarpdağı Y, Okuyar M, et al. Segmentation and classification of skin burn images with artificial intelligence: Development of a mobile application. Burns. 2024;50(4):966–79. 10.1016/j.burns.2024.01.007

51. Chang CW, Lai F, Christian M, et al. Deep learning–assisted burn wound diagnosis: Diagnostic model development Study. JMIR Med Inform. 2021;9(12). 10.2196/22798

52. Chang CW, Wang H, Lai F, et al. Comparison of 3D and 2D area measurement of acute burn wounds with LiDAR technique and deep learning model. Front Artif Intell. 2025;8. 10.3389/frai.2025.1510905

53. Lee S, Rahul, Lukan J, et al. A deep learning model for burn depth classification using ultrasound imaging. J Mech Behav Biomed Mater. 2022;125. 10.1016/j.jmbbm.2021.104930

54. Li Z, Huang J, Tong X, et al. GL-FusionNet: Fusing global and local features to classify deep and superficial partial thickness burn. Math Biosci Eng. 2023;20(6):10153–73. 10.3934/mbe.2023445

